# Optimal protocol design for diffusion-weighted imaging of the prostate: an estimation theory examination of parameter estimate variance

**DOI:** 10.1101/2022.02.26.22271561

**Authors:** Christopher C Conlin, Tyler M Seibert, Anders M Dale

## Abstract

Diffusion-weighted imaging (DWI) is routinely used to aid in the detection and characterization of prostate cancer. Given imaging time constraints in a clinical setting, it is important to maximize the statistical efficiency of a DWI examination of the prostate. The objective is to maximize the accuracy with which microstructural information about the prostate can be obtained while minimizing diffusion scan time.

In this study, we apply estimation theory to evaluate the statistical efficiency of different DWI acquisitions and methods. Specifically, we show that the variance of DWI parameters estimated using nonlinear multiexponential signal models is considerably higher than the variance observed using linear signal models. We then derive a simple analytical expression for the efficiency of a linear estimator and use it to optimize *b*-value sampling for DWI of the prostate.

## Introduction

Diffusion-weighted imaging (DWI) is routinely used to aid in the detection and characterization of prostate cancer (1). While conventional DWI remains largely limited to assessment of relative signal intensity and monoexponential apparent diffusion coefficient (ADC), sophisticated multicompartmental modeling techniques better quantify physiologically meaningful parameters from DWI data (2–6). Compared to conventional DWI, these multicompartmental approaches require more scan time per patient in order to obtain measurements at additional *b*-values and/or echo times (TEs). Given imaging time constraints in a clinical setting, it is important to maximize the statistical efficiency of a DWI examination of the prostate. The objective is to maximize the accuracy with which microstructural information about the prostate can be obtained while minimizing diffusion scan time.

## Methods

### DWI signal modeling

The DWI signal is generally modeled as the sum of individual signal contributions from distinct tissue compartments:

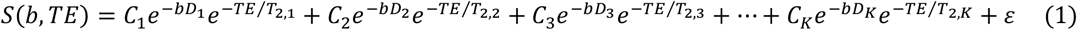

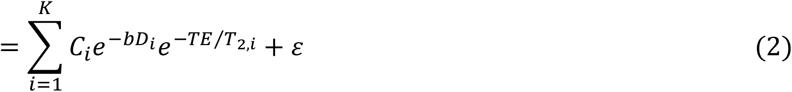

Each compartment is characterized by a linear weight (C), diffusion coefficient (*D*), and transverse relaxation time (*T*_*2*_). The linear weights indicate the relative contributions of each compartment to the overall observed signal. When the DWI signal is normalized, these weights sum to unity and are commonly referred to as the compartmental volume fractions (4,7). The diffusion coefficients and *T*_*2*_ values describe how compartmental signal decays with increasing *b*-value and TE, respectively. Additive measurement noise is represented by *ε*. If the diffusion coefficients and *T*_*2*_ relaxation constants are assumed to be fixed for each compartment, the signal model becomes linear and can be expressed in matrix form as:

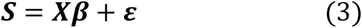

where ***X***is the experimental design matrix that incorporates the exponential decay terms for each compartment at each *b*-value and TE, and ***β***is the vector of linear weights *C* for each compartment.

### Computing estimator variance

An estimator attempts to determine unknown parameters from measured empirical data (8). In the context of the general DWI model presented above, an estimator would aim to determine compartmental *C, D*, and *T*_*2*_ values from DWI signal measurements obtained at different *b*-values and TEs. Estimator variance is an important metric for comparing unbiased estimators, with lower variance indicating a more optimal estimator with higher expected accuracy (9,10). The Cramer-Rao lower bound on the variance of an estimator is given by the inverse of the Hessian matrix of the negative log-likelihood function (11,12). Assuming Gaussian noise, the observed DWI signal measurements may be considered a Gaussian random vector 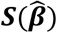 with mean ***S***^**\***^**(*β*)**and covariance **Σ**, where **β** is the concatenation of the various compartmental parameters (*C*_*i*_, *D*_*i*_, *T*_*2,i*_) of the DWI model, and the hat symbol indicates an estimated value. For such a random vector, the joint probability density function (PDF) is given by:

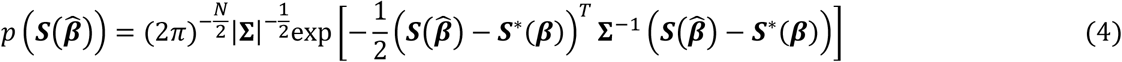

Which yields the negative log likelihood function:

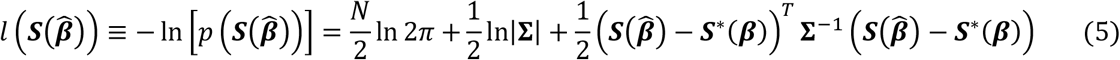

The Hessian matrix of this function describes its curvature in parameter space, and the inverse of the Hessian is the covariance matrix of the uncertain parameters 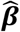. The diagonal elements of the covariance matrix are the variances of each DWI model parameter for a given estimator (11).

### Evaluating parameter variances for nonlinear and linear estimators

Using this numerical approach, we computed parameter variances for a simulated DWI acquisition for one nonlinear and one linear estimator. For both estimators, a 4-compartment signal model was assumed (K = 4 in equation 2) (5). The true DWI model parameters were chosen to reflect values previously reported for prostate tissue (3–5) and are listed in Table 1. For the nonlinear estimator, the nonlinear parameters of the model (compartmental diffusion coefficients, *D*_*i*_, and *T*_*2,i*_ values) were considered free parameters to be estimated from the DWI data, in addition to the linear parameters (compartmental linear weights, *C*_*i*_). For the linear estimator, the nonlinear parameters were considered fixed for each compartment, and only the compartmental linear weights (*C*_*i*_) were estimated from the DWI data. The simulated acquisition protocol was designed to emulate a rigorous examination of the prostate in a research setting, with relatively dense *b*-value and echo time (TE) sampling compared to routine clinical scans. This was done to allow exploration of possibilities, even some that are not presently practical in a clinical setting. Specifically, the simulation acquisition protocol consisted of 5 *b*-values (0, 50, 800, 1500, 3000 s/mm^2^), repeated at 4 different TEs (40, 80, 100, 200 ms). The Hessian matrix of equation 5 was computed numerically, so the nonlinear parameters *D*_*i*_ and *T*_*2,i*_ were log-transformed to improve the stability of numerical differentiation.

**Table 1:** DWI signal model parameters used for simulating prostate tissue.

|  | Linear weights |  |  |  | Diffusion coefficients (mm<sup>2</sup>/s) |  |  |  | T<sub>2</sub> relaxation constants (ms) |  |  |  |
| --- | --- | --- | --- | --- | --- | --- | --- | --- | --- | --- | --- | --- |
| Model parameter | C<sub>1</sub><br>(β<sub>1</sub>) | C<sub>2</sub><br>(β<sub>2</sub>) | C<sub>3</sub><br>(β<sub>3</sub>) | C<sub>4</sub><br>(β<sub>4</sub>) | D<sub>1</sub> (β<sub>5</sub>) | D<sub>2</sub> (β<sub>6</sub>) | D<sub>3</sub> (β<sub>7</sub>) | D<sub>4</sub> (β<sub>8</sub>) | T<sub>2,1</sub><br>(β<sub>9</sub>) | T<sub>2,2</sub><br>(β<sub>10</sub>) | T<sub>2,3</sub><br>(β<sub>11</sub>) | T<sub>2,4</sub><br>(β<sub>12</sub>) |
| True value | 0.3 | 0.4 | 0.2 | 0.1 | 1.0e-4 | 1.8e-3 | 3.6e-3 | 1.0e-2 | 40 | 70 | 100 | 275 |

### Linear estimator efficiency

The statistical efficiency of an unbiased estimator is defined as the reciprocal of the parameter variance per unit of scan time (9,10):

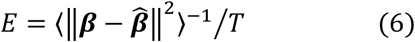

where ⟨·⟩ is the expectation operator, and *T* is the time required for the acquisition. If the estimator under consideration is linear, the numerical Hessian-based approach can be replaced by a simple analytic expression for parameter variance (9,10).

The ordinary least-squares (OLS) solution to equation 3 is given by:

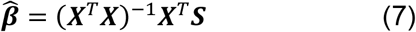

Substituting this OLS solution into equation 6 yields:

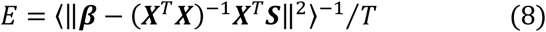

Which can be combined with the expression for DWI signal in equation 3 and simplified to obtain:

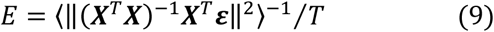

Since ***ε*** is a zero-mean Gaussian random variable, we arrive at the following analytical expression for estimator efficiency for the linear estimator:

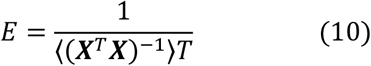

Estimator efficiency *E* provides an objective criterion for evaluating and optimizing the relative expected accuracy of parameter estimates from different DWI acquisition protocols. The **(*X***^*T*^***X*)**^**−1**^ term in equation 10 is the covariance matrix of the parameters, identical to the inverse of the Hessian matrix of the negative log-likelihood function, but simpler to compute in general.

### Optimizing *b*-value sampling to maximize linear estimator efficiency

Using equation 10, an optimal *b*-value sampling scheme was determined for estimating the compartmental diffusion coefficients (*D*_*i*_) of simulated prostate tissue modeled by the parameters listed in Table 1. For a range of maximum *b*-values (from 1000 to 6000 s/mm^2^), a bounded simplex search method (13) was used to compute optimal nonzero *b*-values that maximized the protocol efficiency *E*. Three nonzero *b*-values were assumed initially (along with a *b*=0 s/mm^2^ acquisition), to ensure that the rank of matrix ***X***is at least 4 (the number of parameters to be estimated) but the optimization process was also performed for higher numbers of nonzero b-values (up to 8 in total). Acquisition time *T* was taken to be the minimum repetition time (TR) achievable for the maximum *b*-value on a Discovery MR750 MRI scanner (GE Healthcare, Waukesha, WI) multiplied by the total number of *b*-values. Relative *E* values were computed by normalizing the *E* value obtained for each maximum *b*-value by the maximum *E* value obtained overall. For the protocol with three nonzero *b*-values, sensitivity analysis was performed to examine how varying each *b*-value away from its optimum decreases protocol efficiency. To examine how the value of the diffusion coefficient to be estimated affects the protocol efficiency, the protocol optimization procedure was repeated for three different values of *D*_*1*_: 1e-6, 1e-4, and 1e-3 mm^2^/s.

## Results

The variance of each DWI model parameter is listed in Table 2 for the nonlinear and linear estimators.

**Table 2:** Parameter variances for a simulated DWI acquisition for one nonlinear and one linear estimator. The last row of the table lists the ratio of the nonlinear variance (Var_NL_) over the linear variance (Var_Lin_) for the parameters common to both estimators (the linear weights).

|  | Linear weights |  |  |  | Diffusion coefficients |  |  |  | Transverse relaxation times |  |  |  |
| --- | --- | --- | --- | --- | --- | --- | --- | --- | --- | --- | --- | --- |
| | $C_1$<br>($\beta_1$) | $C_2$ ($\beta_2$) | $C_3$ ($\beta_3$) | $C_4$<br>($\beta_4$) | $\log(D_1)$<br>($\beta_5$) | $\log(D_2)$<br>($\beta_6$) | $\log(D_3)$<br>($\beta_7$) | $\log(D_4)$<br>($\beta_8$) | $\log(T_{2,1})$<br>($\beta_9$) | $\log(T_{2,2})$<br>($\beta_{10}$) | $\log(T_{2,3})$<br>($\beta_{11}$) | $\log(T_{2,4})$<br>($\beta_{12}$) |
| Nonlinear | 259 | 42,616 | 51,249 | 633 | 26,306 | 62,658 | 97,491 | 42,642 | 572 | 22,056 | 99,409 | 54,673 |
| Linear | 8 | 169 | 179 | 11 | N/A (fixed values) |  |  |  | N/A (fixed values) |  |  |  |
| $\text{Var}_{\text{NL}} / \text{Var}_{\text{Lin}}$ | 34 | 252 | 286 | 55 | N/A | | | | N/A | | | |

Figure 1 plots the efficiency of the linear estimator across a range of maximum *b*-values. For the prostate tissue simulated by the parameters in Table 1, the most efficient acquisition protocol having 3 nonzero b-values was achieved using a maximum *b*-value of 2500 s/mm^2^, with optimal nonzero *b*-values of 100, 600, and 2500 s/mm^2^. The sensitivity analysis shown in Figure 2 illustrates how protocol efficiency decreases as each of these optimal *b*-values is varied.

**Figure 1:**
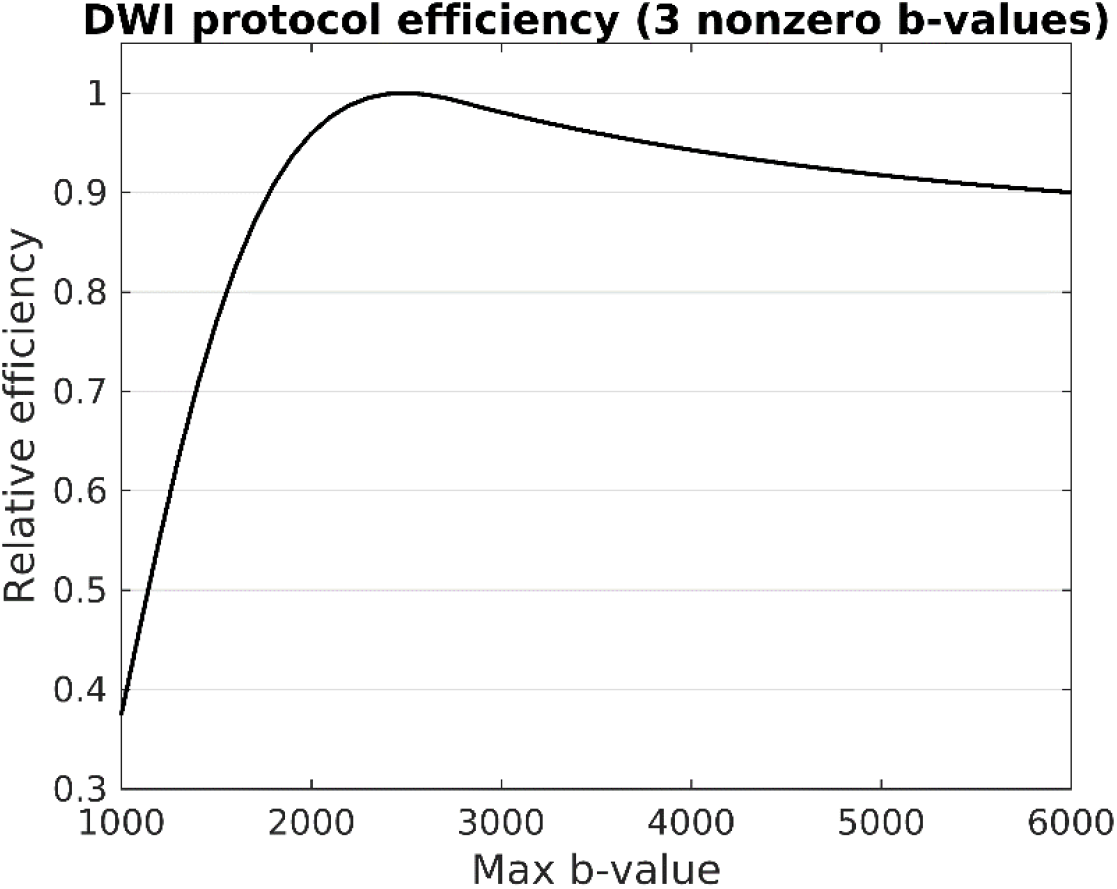
DWI protocol efficiency as a function of the maximum *b*-value acquired. For the simulated prostate tissue to be examined (described by the parameters in Table 1), linear parameter estimation was most efficient for an acquisition protocol with a maximum *b*-value of 2500 s/mm^2^. Optimal nonzero *b*-values for this protocol were 100, 600, and 2500 s/mm^2^.

**Figure 2:**
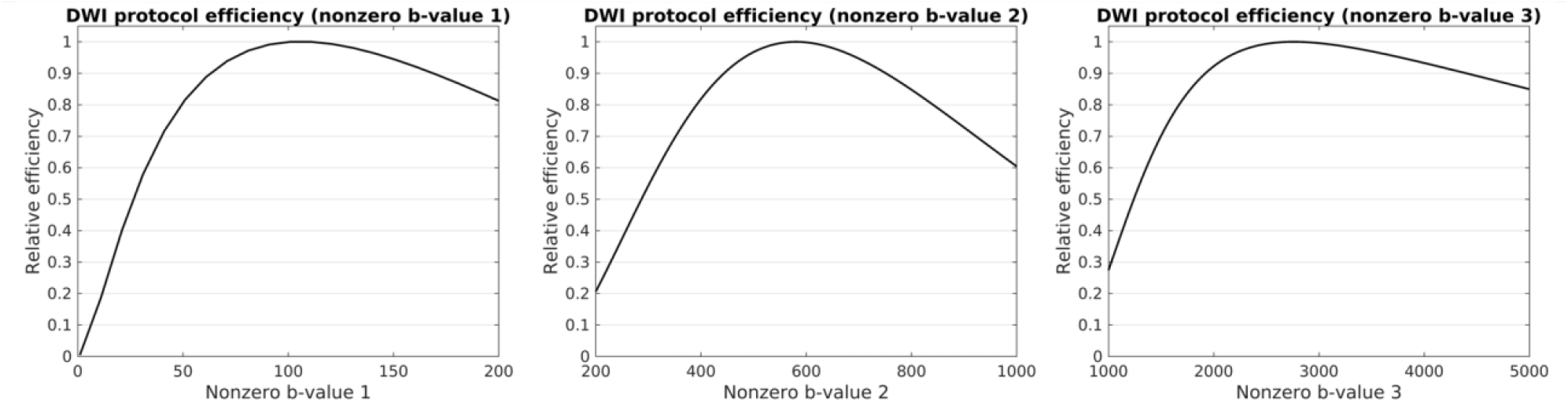
Sensitivity analysis showing how DWI protocol efficiency decreases as each of the three nonzero *b*-values are changed from their optimal values. Optimal nonzero *b*-values for this protocol were 100, 600, and 2500 s/mm^2^.

Allowing for more *b*-values did not result in optimal protocols with additional unique *b*-values, but rather repeated sampling of these initial 3 values (Table 3).

**Table 3:**
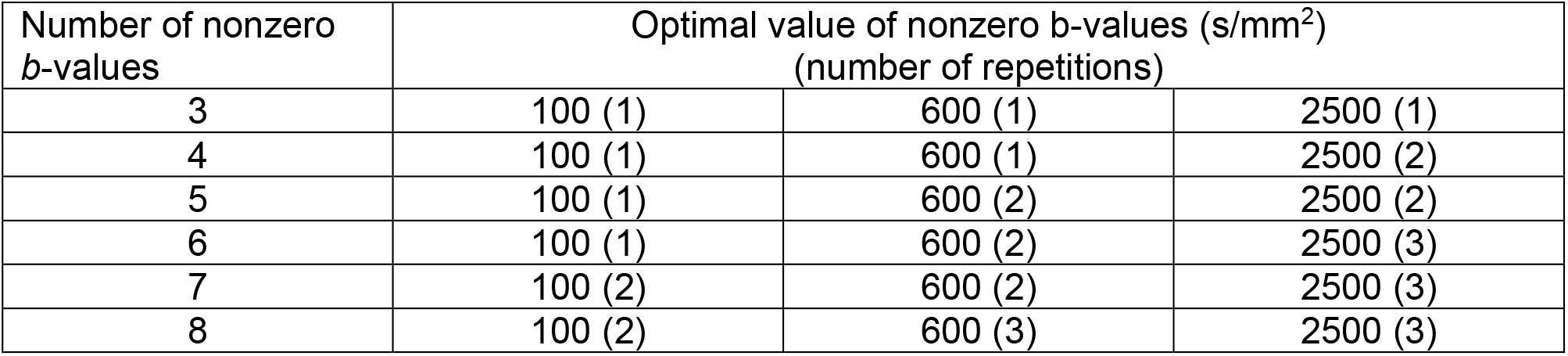
Optimal *b*-values for DWI protocols using different numbers of nonzero *b*-values. A *b*=0 s/mm^2^ acquisition was assumed for each protocol.

Figure 3 demonstrates how DWI estimator efficiency changes based on the underlying diffusion coefficients to be measured. Specifically, as the diffusion coefficient to be estimated decreases, the optimal maximum *b*-value increases.

**Figure 3:**
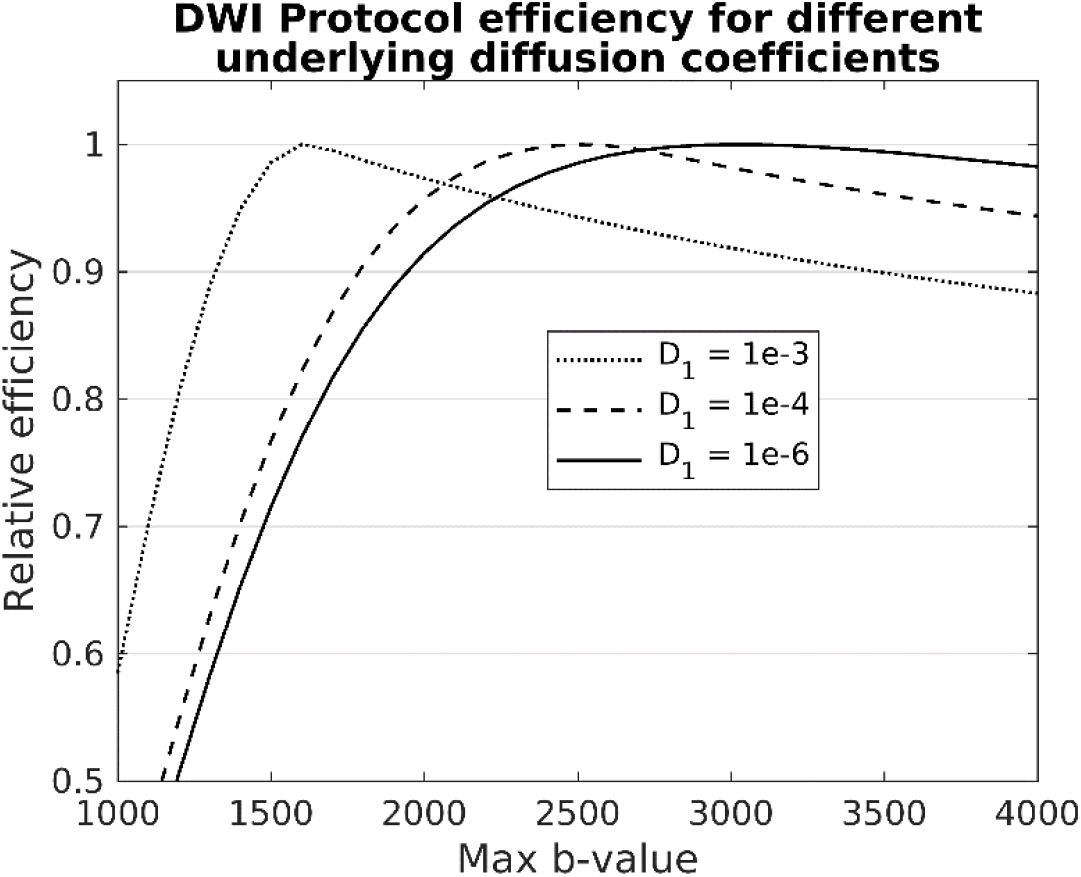
DWI protocol efficiency for different underlying diffusion coefficients. Lower diffusion coefficients are more optimally estimated using higher *b*-values.

## Discussion

Despite the simulated acquisition protocol employed in this study assuming many more measurements than would be realistic in a clinical setting (4 nonzero *b*-values repeated at 4 different TEs), the nonlinear estimator variances were still too large for meaningful determination of the model parameters for the full nonlinear model (Table 2). Such large variances observed for the nonlinear estimator illustrate the inherent difficulty of obtaining accurate parameter estimates from multiexponential fitting of DWI data. Indeed, multiexponential fitting of MRI data in general is often an under-determined or ill-conditioned problem due to limitations on imaging time, leading to solutions that suffer from non-uniqueness, noise amplification, and instability (14,15).

A number of approaches are commonly used to overcome this problem and obtain meaningful parameter estimates from multiexponential fitting of DWI data. Restriction spectrum imaging (RSI) addresses the problem by decoupling the estimation of the nonlinear model parameters (*D*_*i*_, *T*_*2,i*_) from the linear weights (*C*_*i*_). The nonlinear parameters are determined via global fitting to measurements from a large number of voxels simultaneously, and are then fixed per compartment to the globally optimal values (as opposed to being free parameters that are fit voxel-by-voxel) (5). This global fitting process reduces the variance of the nonlinear parameters in proportion to the number of measurements, so they can be reliably estimated for a large enough sample size. In Conlin *et al*., for example, signal measurements from over 200,000 voxels were used to optimize the diffusion coefficients of a 4-compartment DWI signal model of the prostate (5). Once the nonlinear parameters are fixed per compartment, the linear weights can be determined per voxel using linear estimation. As highlighted by Table 2, linear estimation of these parameters has many times lower variance than can be achieved with nonlinear estimation, making voxel-wise measurements of *C* much more accurate. Other DWI techniques like intravoxel incoherent motion (IVIM) imaging (17), Vascular, Extracellular, and Restricted Diffusion for Cytometry in Tumors (VERDICT) MRI (2), and Hybrid Multidimensional MRI (4) endeavor to fit the nonlinear model parameters on a voxel-wise basis. To overcome the large variances inherent in the estimation of these parameters, bounds are generally imposed on the range of possible values they can take during the nonlinear fitting process (2,4,18). This approach is analogous to the nonlinear parameter fixing in RSI, but permits limited variation in the parameter estimates between voxels. The nonlinear parameter estimates may also be analyzed after averaging across all voxels in a chosen region of interest (ROI) or tissue to further reduce estimation variance (19–21).

Equation 10 provides an objective metric of linear estimator efficiency that can readily be used to optimize DWI acquisition protocols. For a 4-compartment model of prostate tissue with diffusion coefficients of 1.0e-4, 1.8e-3, 3.6e-3, and 1.0e-2 mm^2^/s, the optimal nonzero *b*-values which maximized estimator efficiency were calculated to be 100, 600, and 2500 s/mm^2^. As shown in Table 3, repeated measurements at these optimal *b*-values was more efficient than including additional unique *b*-values. This demonstrates that dense protocols with many evenly spaced *b*-values are not generally the optimal approach for DWI experiments. While tissues characterized by different diffusion coefficients will have different optimal *b*-values, the proposed strategy for protocol optimization still applies; the only change needed is to recompute the constant terms of the design matrix ***X***using the new diffusion coefficients. In general, smaller diffusion coefficients are more optimally estimated by higher *b*-values (Figure 3). This finding is particularly important in the context of prostate cancer imaging, where tumors are characterized by increased cellularity and more restricted diffusion (2,22,23). The optimal maximum b-value reported in this study (2500 s/mm^2^) is higher than what is typically prescribed for prostate DWI clinically (24), suggesting that reevaluation of current clinical guidelines may be warranted.

## Data Availability

All data produced in the present study are available upon reasonable request to the authors

## References

1. Turkbey B, Rosenkrantz AB, Haider MA, et al. Prostate Imaging Reporting and Data System Version 2.1: 2019 Update of Prostate Imaging Reporting and Data System Version 2. European Urology. 2019;76(3):340–351. doi: 10.1016/j.eururo.2019.02.033.

2. Panagiotaki E, Chan RW, Dikaios N, et al. Microstructural Characterization of Normal and Malignant Human Prostate Tissue With Vascular, Extracellular, and Restricted Diffusion for Cytometry in Tumours Magnetic Resonance Imaging. Investigative Radiology. 2015;50(4):218. doi: 10.1097/RLI.0000000000000115.

3. Hectors SJ, Said D, Gnerre J, Tewari A, Taouli B. Luminal Water Imaging: Comparison With Diffusion-Weighted Imaging (DWI) and PI-RADS for Characterization of Prostate Cancer Aggressiveness. Journal of Magnetic Resonance Imaging. 2020;52(1):271–279. doi: 10.1002/jmri.27050.

4. Chatterjee A, Bourne RM, Wang S, et al. Diagnosis of Prostate Cancer with Noninvasive Estimation of Prostate Tissue Composition by Using Hybrid Multidimensional MR Imaging: A Feasibility Study. Radiology. 2018;287(3):864–873. doi: 10.1148/radiol.2018171130.

5. Conlin CC, Feng CH, Rodriguez-Soto AE, et al. Improved Characterization of Diffusion in Normal and Cancerous Prostate Tissue Through Optimization of Multicompartmental Signal Models. Journal of Magnetic Resonance Imaging. 2021;53(2):628–639. doi: https://doi.org/10.1002/jmri.27393.

6. Feng CH, Conlin CC, Batra K, et al. Voxel-level Classification of Prostate Cancer on Magnetic Resonance Imaging: Improving Accuracy Using Four-Compartment Restriction Spectrum Imaging. Journal of Magnetic Resonance Imaging. 2021;54(3):975–984. doi: 10.1002/jmri.27623.

7. Panagiotaki E, Schneider T, Siow B, Hall MG, Lythgoe MF, Alexander DC. Compartment models of the diffusion MR signal in brain white matter: A taxonomy and comparison. NeuroImage. 2012;59(3):2241–2254. doi: 10.1016/j.neuroimage.2011.09.081.

8. Walter E, Pronzato L. Identification of Parametric Models: From Experimental Data. Springer; 1997.

9. Dale AM. Optimal experimental design for event-related fMRI. Human Brain Mapping. 1999;8(2–3):109–114. doi: 10.1002/(SICI)1097-0193(1999)8:2/3<109::AID-HBM7>3.0.CO;2-W.

10. White NS, Dale AM. Optimal diffusion MRI acquisition for fiber orientation density estimation: An analytic approach. Human Brain Mapping. 2009;30(11):3696–3703. doi: 10.1002/hbm.20799.

11. Appendix A: Relationship between the Hessian and Covariance Matrix for Gaussian Random Variables. Bayesian Methods for Structural Dynamics and Civil Engineering. John Wiley & Sons, Ltd; 2010. p. 257–262. doi: 10.1002/9780470824566.app1.

12. Kay SM. Chapter 3: Cramer-Rao Lower Bound. Fundamentals of Statistical Signal Processing, Volume I: Estimation Theory. 1st edition. Pearson; 1993.

13. Lagarias JC, Reeds JA, Wright MH, Wright PE. Convergence Properties of the Nelder--Mead Simplex Method in Low Dimensions. SIAM J Optim. 1998;9(1):112–147. doi: 10.1137/S1052623496303470.

14. Raj A, Pandya S, Shen X, LoCastro E, Nguyen TD, Gauthier SA. Multi-Compartment T2 Relaxometry Using a Spatially Constrained Multi-Gaussian Model. PLOS ONE. Public Library of Science; 2014;9(6):e98391. doi: 10.1371/journal.pone.0098391.

15. Graham SJ, Stanchev PL, Bronskill MJ. Criteria for analysis of multicomponent tissue T2 relaxation data. Magnetic Resonance in Medicine. 1996;35(3):370–378. doi: 10.1002/mrm.1910350315.

16. Tikhonov AN, Goncharsky AV, Stepanov VV, Yagola AG. Regularization methods. In: Tikhonov AN, Goncharsky AV, Stepanov VV, Yagola AG, editors. Numerical Methods for the Solution of Ill-Posed Problems. Dordrecht: Springer Netherlands; 1995. p. 7–63. doi: 10.1007/978-94-015-8480-7_2.

17. Le Bihan D, Breton E, Lallemand D, Aubin ML, Vignaud J, Laval-Jeantet M. Separation of diffusion and perfusion in intravoxel incoherent motion MR imaging. Radiology. 1988;168(2):497–505. doi: 10.1148/radiology.168.2.3393671.

18. Döpfert J, Lemke A, Weidner A, Schad LR. Investigation of prostate cancer using diffusion-weighted intravoxel incoherent motion imaging. Magnetic Resonance Imaging. 2011;29(8):1053–1058. doi: 10.1016/j.mri.2011.06.001.

19. Andreassen MMS, Rodríguez-Soto AE, Conlin CC, et al. Discrimination of Breast Cancer from Healthy Breast Tissue Using a Three-component Diffusion-weighted MRI Model. Clin Cancer Res. American Association for Cancer Research; 2021;27(4):1094–1104. doi: 10.1158/1078-0432.CCR-20-2017.

20. Donati OF, Mazaheri Y, Afaq A, et al. Prostate Cancer Aggressiveness: Assessment with Whole-Lesion Histogram Analysis of the Apparent Diffusion Coefficient. Radiology. 2013;271(1):143–152. doi: 10.1148/radiol.13130973.

21. Vargas HA, Akin O, Franiel T, et al. Diffusion-weighted Endorectal MR Imaging at 3 T for Prostate Cancer: Tumor Detection and Assessment of Aggressiveness. Radiology. 2011;259(3):775–784. doi: 10.1148/radiol.11102066.

22. Liss MA, White NS, Parsons JK, et al. MRI-Derived Restriction Spectrum Imaging Cellularity Index is Associated with High Grade Prostate Cancer on Radical Prostatectomy Specimens. Front Oncol. 2015;5. doi: 10.3389/fonc.2015.00030.

23. Kuwano H, Miyazaki T, Tsutsumi S, et al. Cell Density Modulates the Metastatic Aggressiveness of a Mouse Colon Cancer Cell Line, Colon 26. OCL. 2004;67(5–6):441–449. doi: 10.1159/000082929.

24. Weinreb JC, Barentsz JO, Choyke PL, et al. PI-RADS Prostate Imaging – Reporting and Data System: 2015, Version 2. European Urology. 2016;69(1):16–40. doi: 10.1016/j.eururo.2015.08.052.

